# Prediction of Severe Obstructive Coronary Artery Disease Using Radiomic Features from Resting Cardiac Ultrasound Images: A Pilot Study

**DOI:** 10.1101/2024.03.28.24305048

**Authors:** Daniel Aziz, Ankush D. Jamthikar, Abhijit Bhattaru, Karthik Desingu, Nivedita Rajiv, Kameswari Maganti, Yasmin Hamirani, Sabahat Bokhari, Naveena Yanamala, Partho P. Sengupta

## Abstract

**Background:** Although cardiac ultrasound is frequently performed in patients with chest pain, the probability of obstructive coronary artery disease (CAD) cannot be quantified. We investigated the potential of cardiac ultrasound radiomics (ultrasomics) to identify obstructive CAD using limited echocardiography frames, suitable for cardiac point-of-care ultrasound evaluation.

**Methods:** In total, 333 patients who were either healthy controls (n=30), undergoing invasive coronary procedures (n=113), or coronary CT angiography (n=190) were divided into two temporally distinct training (n=271) and testing (n=62) cohorts. Machine learning models were developed using ultrasomics for predicting severe CAD (stenosis >70%) and compared with regional LV wall motion abnormalities (RWMA).

**Results:** In total, 94 (28.2%) patients had severe CAD with 50 (15.0%) having high-risk CAD defined as left main stenosis >50% (n=11), multivessel CAD (n=43), or 100% occlusion (n=20). The ultrasomics model was superior to RWMA for predicting severe CAD [area under the receiver operating curve (AUC) of 0.80 (95% confidence interval [CI]: 0.74 to 0.86) vs. 0.67 (95% CI: 0.61-0.72), p=0.0014] in the training set and [0.77 (95% CI: 0.64-0.90) vs. 0.70 (95% CI: 0.56-0.81), p=0.24] in the test set, respectively. The model also predicted high-risk CAD with an AUC of 0.84 (95% CI: 0.77-0.90) in the training set and 0.70 (95% CI: 0.48-0.88) in the test set. A combination of ultrasomics with RWMA showed incremental value over RWMA alone for predicting severe CAD.

**Conclusions:** Cardiac ultrasomic features extracted from limited echocardiography views can aid the development of machine learning models to predict the presence of severe obstructive CAD.

## Introduction

Coronary artery disease (CAD) remains one of the leading causes of global morbidity and mortality, with approximately 9 million annual deaths annually, a prevalence of 190 million individuals, and minimal improvement in incidence since 2008 (1,2). Clinical presentations range from stable angina to acute coronary syndromes due to luminal narrowing or thrombi formation.

Recent guidelines have given a Class I recommendation for coronary computed tomography angiography (CCTA) in patients with suspected CAD who are low or intermediate risk while recommending invasive coronary angiography for patients with unstable coronary syndromes or those who cannot be medically managed and are at high risk for adverse cardiac events (3,4). To standardize patient management and severity assessment, the Coronary Artery Disease – Reporting and Data System (CAD-RADS) was introduced, grading luminal stenosis from absent (category 0) to total occlusion (category 5) (5). Elevated CAD-RADS categories, specifically over 4 (>70% luminal stenosis) —referred to as severe obstructive CAD – correlate unequivocally with an increased adverse risk for unfavorable outcomes (6). Despite this increased risk, there remains an unmet need for safe, non-invasive, and readily available algorithms that can guide triaging of patients with suspected severe obstructive CAD.

The implementation of artificial intelligence (AI) in image analysis has facilitated the automation of tasks that traditionally relied on expert input. Specifically, the combination of AI and ‘omics’-based methods offers the potential for capturing individualized human health complexities. ’Radiomics’, the extraction of quantitative pixel-level data from medical images, has been applied extensively in cancer and neuroscience research (7). In cardiology, radiomics has been utilized in cardiac computed tomography (CT) and magnetic resonance imaging (CMR); yet its role in extracting cardiac ultrasound textural properties remains largely unexplored (8).

In our recent work, we demonstrated the potential of cardiac ultrasound radiomics (ultrasomics) in capturing subtle LV morphological and functional changes that may not be apparent on visual inspection alone (9). Cardiac ultrasound imaging is frequently performed in patients with chest pain to assess global and regional left ventricular function and rule out alternative diagnoses. For the present investigation, we therefore explored the integration of ultrasomics with machine learning to predict the presence of severe obstructive CAD from limited echocardiography views which are suitable for cardiac point-of-care ultrasound evaluation.

## Methods

### Study Population

We included 333 participants across three centers to establish training and validation cohorts: a) patients from a previously published prospective study (n=105) that enrolled consecutive patients undergoing clinically indicated coronary angiogram (n=75) and healthy controls (n=30) at West Virginia University, Morgantown (WVU), WV, USA (10), b) a previously published cohort who underwent CCTA in ambulatory settings at Mount Sinai Medical center (MSM), New York (n= 190) (11), and c) a retrospective cohort of non-ST elevation myocardial infarction (NSTEMI) patients undergoing invasive coronary angiogram at Robert Wood Johnson University Hospital (RWJUH) (n=38). The inclusion and exclusion criteria for patients prospectively enrolled at WVU and MSM have been previously published (10,11). The institutional ethics committee of Rutgers Robert Wood Johnson Medical School and RWJUH approved the study to enable the analysis of the clinical, echocardiography, coronary CT, and invasive coronary angiography data.

The NSTEMI patients selected at RWJUH were specifically chosen to ensure diversity in the patient population and to evaluate how well the machine learning model can predict severe obstructive disease in CAD patients who were not well-represented in previous cohorts. Exclusion criteria for patients at RWJUH included (1) patients discharged to institutionalized care, (2) type 2-5 acute myocardial infarction (AMI), (3) co-existing terminal illness, such as cancer, (4) alternative diagnosis for elevated cardiac troponin values (e.g. myocarditis, non-ischemic cardiomyopathies, moderate-severe valvular heart disease), (5) pregnancy, and (6) technically insufficient imaging for one out of 2 views: parasternal long-axis (PLAX) and apical 4 chambers (A4C).

To allow for a realistic evaluation of models on subsequent data, training and test datasets were derived from a temporal split. 80% of the chronologically admitted patients were selected for the training set along with healthy controls (n=271), while the remaining 20% of patients were included in the test set (n=62).

### Clinical Endpoints

Severe obstructive coronary artery disease was defined as left main artery stenosis greater than 50% or stenosis greater than 70% in any other significant vessels such as the left anterior descending, left circumflex, or right coronary artery. Additionally, high-risk coronary artery disease was defined as left main artery stenosis greater than 50%, multi-vessel obstructive CAD (>70%), or the presence of 100% coronary artery disease in any other significant vessel.

### Echocardiography Image Acquisition, Preprocessing, and Semantic Segmentation

Echocardiographic examinations performed at each center using commercial ultrasound systems included 2D, color Doppler, spectral Doppler, and tissue Doppler imaging. All measurements and evaluations were conducted by board-certified echocardiographers. All studies were stored on a cloud-based analysis platform (CoreSound Inc.) and all measurements including RWMA were standardized using a validated vendor-independent algorithm (Us2.ai Inc.) (12).

For the ultrasomics machine-learning pipeline, we employed an automated echocardiography workflow consisting of four stages: preprocessing, echocardiographic view identification, semantic segmentation, and ultrasound feature extraction (Figure 1). To enable the adoption of the models on cardiac point-of-care-ultrasound (POCUS), we used two echocardiography views that are recommended in a set of limited views obtained for cardiac POCUS: parasternal long-axis (PLAX) or apical four-chamber views (A4C) (13). To ensure uniformity and compatibility across data processing, 2D echocardiographic images from diverse formats (e.g., .avi, .mp4 and .dcm) were converted into DICOM using the proprietary Sante DICOM software (SanteSoft, Nicosia, Cyprus, Greece). Images not amenable to ultrasomic analysis, such as those containing Doppler data or dual ultrasound regions primarily used for static measurements, were subsequently filtered out. The view identification stage utilized these processed multi-beat echocardiogram DICOM files to classify the transthoracic echocardiography views into A4C and PLAX views. The semantic segmentation stages involved the standard echoCV algorithm (14), which automatically delineated the left ventricular myocardium region at the end-diastole from the echocardiography recordings corresponding to the PLAX and A4C views. In the PLAX view, the region corresponding to the intra-ventricular septum and the posterior wall were combined to form a combined LV myocardial region. In this study, we modified the existing echoCV platform (14) to be executed using Python 3.2 with CUDA 10.0 support provided by TensorFlow 1.15.0. To maintain uniformity across different image sizes, we resized the segmented images to 1024 by 1024 pixels. Our prior study discussed the use of echoCV and its validation for predicting LV remodeling (15). From the semantic segmentation stages, we received the binary mask for the region of interest for LV myocardium, which was further utilized to extract the ultrasomic features from the original grayscale images.

**Figure 1:**
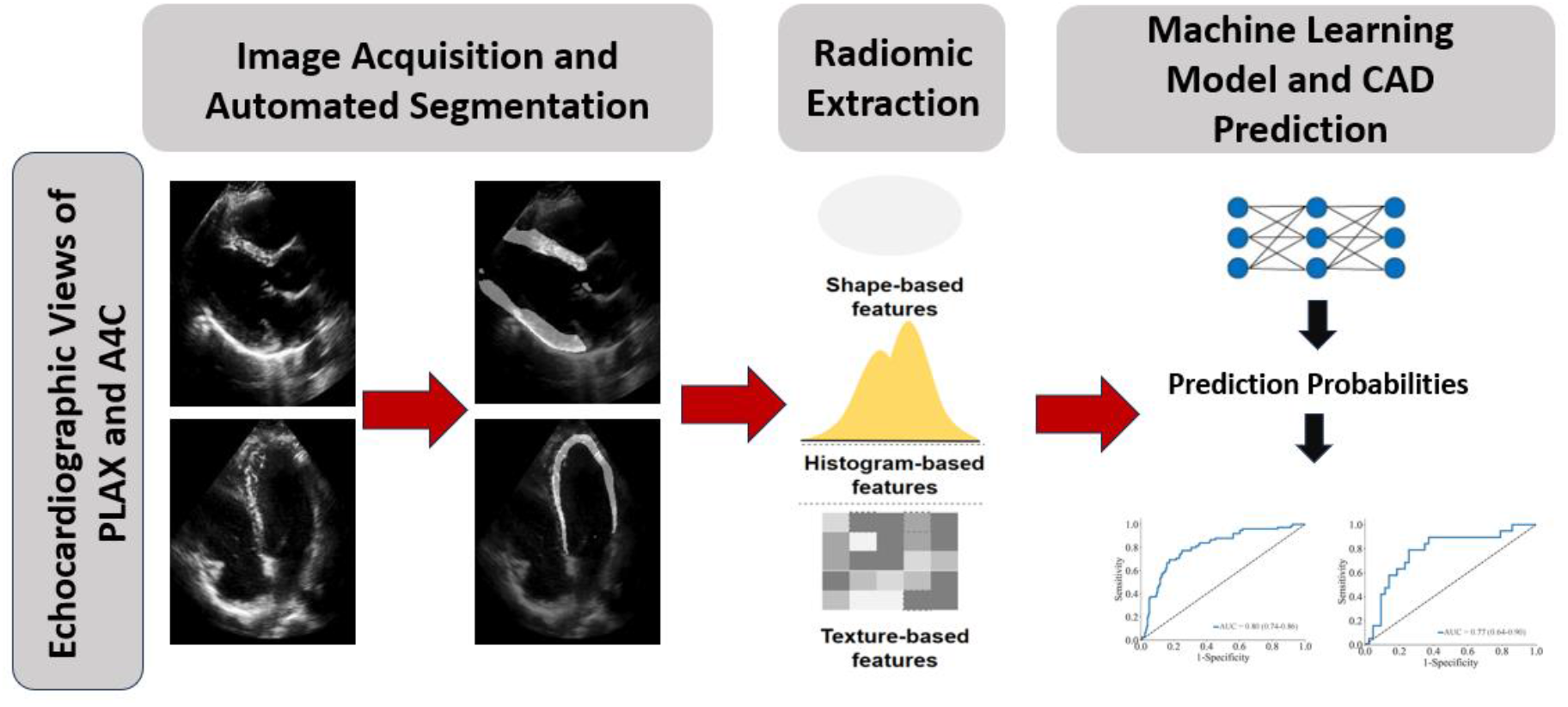
Central Illustration: Depiction of the ultrasomics machine-learning pipeline, including the preprocessing, echocardiographic view identification, semantic segmentation, and ultrasound feature extraction. After extraction, machine learning was employed to create a predictive ultrasomics model for severe CAD.

### Cardiac Ultrasound Ultrasomics Extraction

Ultrasomic features were extracted using PyRadiomics (version 3.0.1, Python Software Foundation [15]) and SimpleITK (version 2.2.0, Insight Software Consortium [16]) within the open-source Python framework (version 3.7.13). A total of 99 ultrasomic features, corresponding to the shape, texture, and intensity distribution within the region of interest, were extracted from the LV myocardium region, resulting in 198 features for both the PLAX and A4C views.

### Machine Learning Model Development

We developed a supervised machine learning (ML)-based model to predict severe, obstructive coronary artery disease using the open-source scikit-learn platform (scikit-learn 1.3.2). To address variability among the data sources, including image storage formats and scanner-related variability, we implemented a two-step normalization approach. First, the ultrasomics features were independently normalized using the cumulative distribution function, ensuring that the data fell within the scale of 0-1 for all features (15). Next, the ultrasomics from each of the three datasets were temporally split into training and test sets. In the second normalization step, we computed standard normalization, also known as z-score normalization, on the temporally split training dataset. The same transformations were applied to the test dataset.

After training and testing feature preparation, we trained a support vector machine (SVM)-based algorithm, utilizing a linear kernel with a C value of 1 as hyperparameters to train the model on the training dataset. To prevent overfitting, a 10-fold cross-validation strategy was employed on the training dataset to evaluate the overall performance of the model. In 10-fold cross-validation, the complete training dataset was split into 10 equal parts. At each of the 10 iterations, the model was trained on nine parts (i.e., 90% of the training data) and evaluated on the remaining part (i.e., 10% of the dataset). The performance of the model was evaluated on the test dataset using five performance evaluation metrics: sensitivity, specificity, F1-score, accuracy, and area under the receiver operating characteristic curve (AUC). The performance on the training dataset is presented as mean ± standard deviation for each of these five performance evaluation metrics. The prediction probabilities from the model were on a 0-1 scale and served as a continuous ultrasomics score representing obstructive coronary artery disease. They can also be interpreted as a binary outcome, with any value ≥ 0.5 indicating the presence of severe CAD, and a value < 0.5 indicating the absence of severe CAD within a patient.

### Performance evaluation and statistical analysis

Baseline characteristics for the training and test datasets are presented as median (interquartile range) for the continuous variables and total count (percentage) for the categorical variables. Continuous variables between training and test datasets were compared using the Mann-Whitney U test, while categorical variables were compared using the Fisher exact test. For categorical values with more than two categories, a chi-square test was used. The presence of regional wall motion abnormalities was reported as a categorical variable.

To calculate the AUC, a DeLong test was used to determine the ability of the model to predict severe and high-risk CAD. DeLong test was also used to calculate the AUC of the presence of RWMA for predicting severe and high-risk CAD. Logistic regression was performed using probabilities from our ultrasomics model for severe CAD, along with the presence of RWMA, to predict severe CAD. High-risk CAD prediction was conducted using ultrasomics model probabilities and the presence of RWMA. The incremental value of our ultrasomics model was compared with the predictive value of the presence of RWMA using a DeLong test to compare ROC curves. All statistical analyses were conducted using MedCalc® Statistical Software version 22.021. All statistical tests were two-sided with a significance level of 0.05 unless otherwise specified.

## Results

### Baseline characteristics

The clinical characteristics of all 333 study participants were divided into training and test sets, as assessed, and presented in Table 1. Similarly, the baseline characteristics for study participants who underwent CT coronary angiography (n=190), and invasive coronary angiography (n=143) were also assessed and presented as training and test sets in Table 2 and Table 3, respectively. Overall, in our training cohort of 271 patients, of whom 88 underwent coronary angiograms, 153 underwent coronary CT angiography, and 30 were healthy controls, a total of 75 (27.7%) subjects had severe CAD. In the testing cohort, 19 out of 62 (30.6%) subjects had severe CAD. In the training cohort, 42 out of 271 patients (15.5%) had high-risk CAD, while in the test cohort, 8 out of 62 patients (12.9%) had the same composite and was not significantly different between the groups. Between the overall training and test cohorts, statistically significant differences included: age (57 vs 63 years old, p=0.0032), prevalence of hypertension (65.3% vs. 83.9%, p=0.00387), and prevalence of smoking (42.4% vs. 56.5%, p=0.0487), respectively. The presence of RWMA was observed in 27 out of 271 patients (10%) in the training set and 6 out of 62 patients (9.7%) in the test set.

**Table 1:**
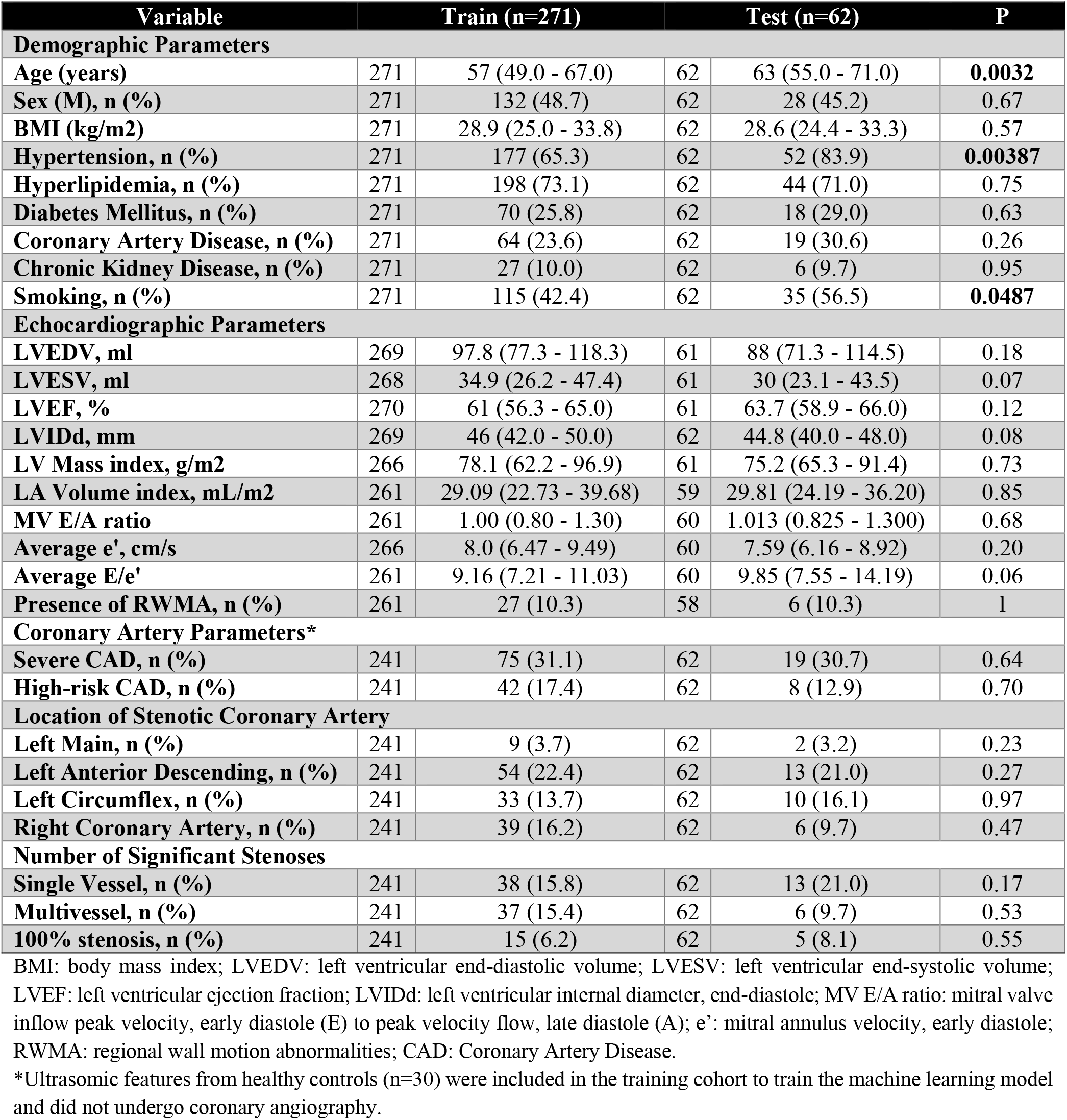
Baseline characteristics for the training and test datasets used to develop and validate the machine learning models.

**Table 2:**
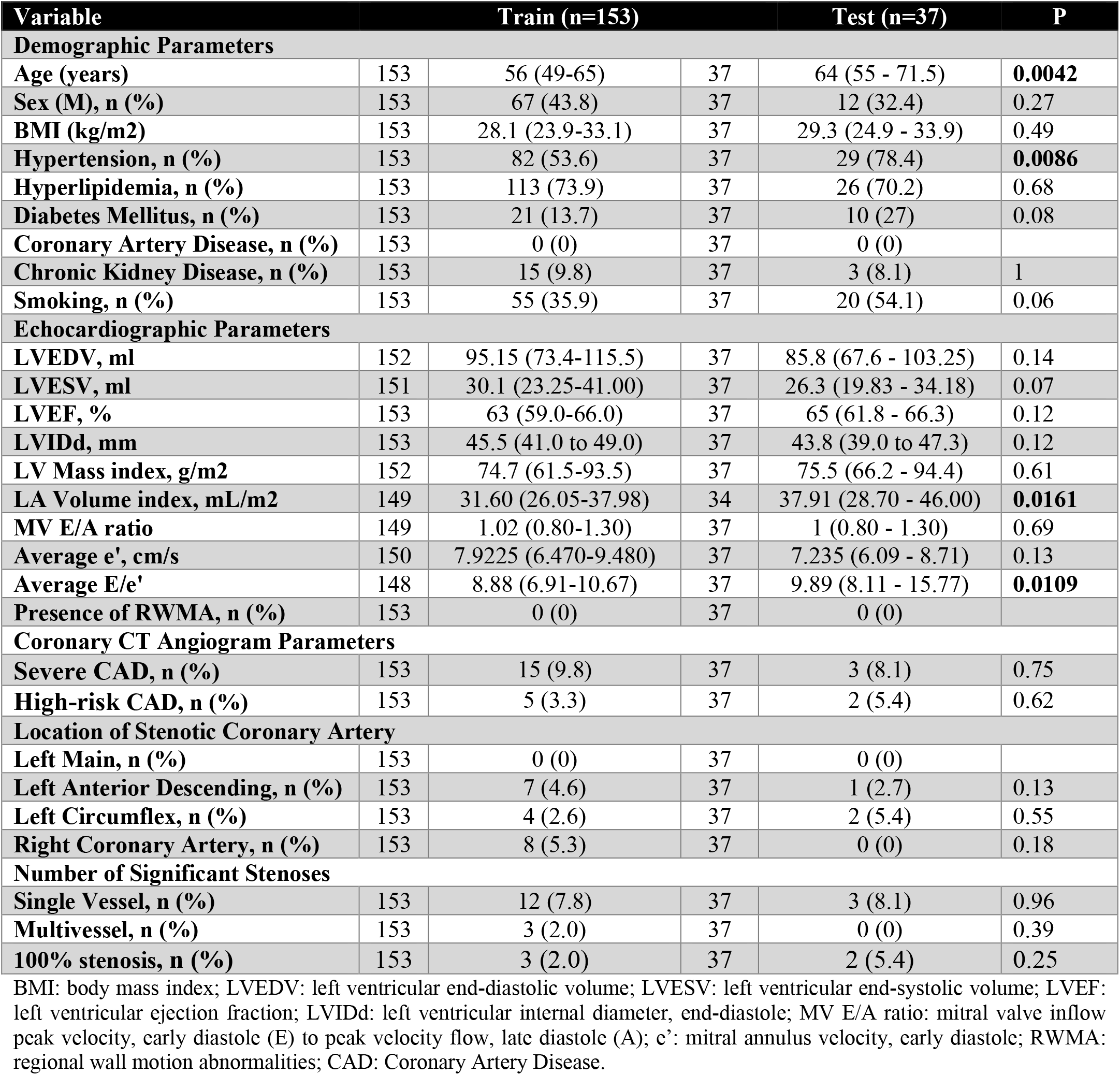
Baseline characteristics for the study participants who underwent coronary CT angiography.

**Table 3:** Baseline characteristics of study participants who underwent invasive coronary angiography.

| Variable | Train (n=118) |  | Test (n=25) |  | P |
| --- | --- | --- | --- | --- | --- |
| Demographic Parameters |  |  |  |  |  |
| Age (years) | 118 | 58.5 (49.0 - 68.0) | 25 | 60.0 (53.7 - 71.0) | 0.22 |
| Sex (M), n (%) | 118 | 65 (55.1) | 25 | 16 (64.0) | 0.51 |
| BMI (kg/m2) | 118 | 29.2 (26.4 to 34.2) | 25 | 27.3 (24.2 - 32.5) | 0.06 |
| Hypertension, n (%) | 118 | 95 (80.5) | 25 | 23 (92.0) | 0.25 |
| Hyperlipidemia, n (%) | 118 | 85 (72.0) | 25 | 18 (72.0) | 1 |
| Diabetes Mellitus, n (%) | 118 | 49 (41.5) | 25 | 8 (32.0) | 0.50 |
| Coronary Artery Disease, n (%) | 118 | 64 (54.2) | 25 | 19 (76.0) | **0.0483** |
| Chronic Kidney Disease, n (%) | 118 | 12 (10.2) | 25 | 3 (12.0) | 0.73 |
| Smoking, n (%) | 118 | 60 (50.8) | 25 | 15 (60) | 0.51 |
| Echocardiographic Parameters |  |  |  |  |  |
| LVEDV, ml | 117 | 99.8 (84.9 to 120.2) | 24 | 102.4 (76.1 to 128.1) | 0.93 |
| LVESV, ml | 117 | 40.5 (29.1 to 55.9) | 24 | 41.5 (28.9 to 52.2) | 0.64 |
| LVEF, % | 117 | 58 (53.7 to 63.9) | 24 | 61.4 (53.2 to 63.9) | 0.51 |
| LVIDd, mm | 116 | 46.0 (42.0 to 50.5) | 25 | 46.0 (41.7 to 48.4) | 0.37 |
| LV Mass index, g/m2 | 114 | 83.4 (63.9 to 103.7) | 25 | 73.3 (63.9 to 81.5) | 0.14 |
| LA Volume index, mL/m2 | 112 | 27.29 (22.80 to 32.73) | 25 | 20.7 (17.61 to 28.13) | **0.0011** |
| MV E/A ratio | 112 | 0.967 (0.812 to 1.286) | 23 | 1.038 (0.89 to 1.45) | 0.24 |
| Average e', cm/s | 116 | 8.0 (6.45 to 9.62) | 23 | 8.0 (6.79 to 9.04) | 0.88 |
| Average E/e' | 113 | 9.55 (7.61 to 12.19) | 23 | 8.74 (7.52 to 11.34) | 0.86 |
| Presence of RWMA, n (%) | 108 | 27 (25) | 21 | 6 (28.6) | 0.79 |
| Invasive Coronary Angiogram Parameters* |  |  |  |  |  |
| Severe CAD, n (%) | 88 | 60 (68.2) | 25 | 16 (64.0) | 0.68 |
| High-risk CAD, n (%) | 88 | 37 (42.0) | 25 | 6 (24.0) | 0.27 |
| Location of Stenotic Coronary Artery |  |  |  |  |  |
| Left Main, n (%) | 88 | 9 (10.2) | 25 | 2 (8.0) | 0.55 |
| Left Anterior Descending, n (%) | 88 | 47 (53.4) | 25 | 12 (48.0) | 0.05 |
| Left Circumflex, n (%) | 88 | 29 (32.9) | 25 | 8 (32.0) | 0.69 |
| Right Coronary Artery, n (%) | 88 | 31 (35.2) | 25 | 6 (24.0) | **0.0143** |
| Number of Significant Stenoses |  |  |  |  |  |
| Single Vessel, n (%) | 88 | 26 (29.5) | 25 | 10 (40.0) | 0.23 |
| Multivessel, n (%) | 88 | 34 (38.6) | 25 | 6 (24.0) | 0.37 |
| 100% stenosis, n (%) | 88 | 12 (13.6) | 25 | 3 (12.0) | 0.98 |

BMI: body mass index; LVEDV: left ventricular end-diastolic volume; LVESV: left ventricular end-systolic volume; LVEF: left ventricular ejection fraction; LVIDd: left ventricular internal diameter, end-diastole; MV E/A ratio: mitral valve inflow peak velocity, early diastole (E) to peak velocity flow, late diastole (A); e': mitral annulus velocity, early diastole; RWMA: regional wall motion abnormalities; CAD: Coronary Artery Disease.
\*Ultrasomic features from healthy controls (n=30) were included in training cohort to train the machine learning model and did not undergo coronary angiography.

Patients undergoing CCTA were also distributed into training (n=153) and test (n=37) cohorts (Table 2) and overall had a lower prevalence of severe CAD in both the training (9.8%) and test (8.1%) cohorts. Patients in the test group were significantly older (56 vs. 64 years old, p=0.0042) and had a higher prevalence of hypertension (53.6% vs. 78.4%, p=0.0086) compared to those in the training set. The distribution of echocardiographic and coronary angiographic findings is shown in Table 2.

In patients who underwent invasive coronary angiography (n=113), there was a higher overall prevalence of severe CAD with 60 out of 88 (68.2%) in the training group and 16 out of 25 (66.7%) in the test group, than the CCTA group (Table 3).

### Predicting Severe Coronary Artery Disease

The ML model was evaluated with 10-fold cross-validation in the training set and demonstrated better average performance overall, with an AUC value (Figure 2) of 0.80 (95% confidence interval [CI]: 0.74 to 0.86). On the test set, the model exhibited equivalent or better predictive performance, with an AUC of 0.77 (95% CI: 0.64-0.90). The sensitivity and specificity of this model on the training and test sets were as follows: sensitivity of 77.3% and specificity of 69.4% in the training cohort, sensitivity of 79% and specificity of 72.1% in the testing cohort. The ultrasomics model had a better diagnostic performance with an AUC of 0.80 (95% CI: 0.74-0.86) than the RWMA assessment with an AUC of 0.67 (95% CI: 0.61-0.72, p=0.0014) in the training set. Similar performance was also observed in the test set with an AUC of 0.77 (95% CI: 0.64-0.90) vs. 0.70 (95% CI: 0.56-0.81, p=0.24).

**Figure 2:**
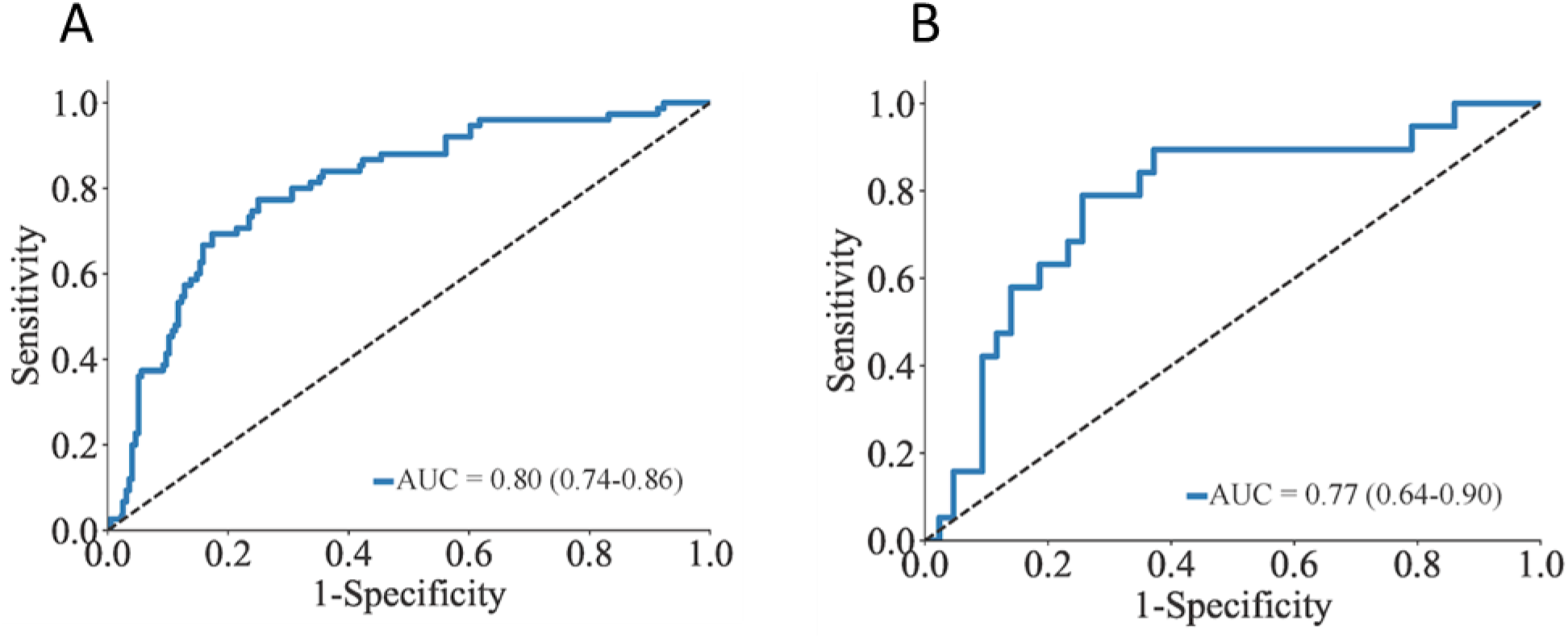
Prediction of Severe CAD From ML Model. (A) Receiver-operating characteristic (ROC) curve of the training cohort with 10-fold cross-validation for predicting severe CAD and (B) ROC curve for the prediction of severe CAD in the validation (test) cohort.

### Predicting High-Risk Coronary Artery Disease

In the training set (Figure 3A), ultrasomics predicted high-risk CAD, with an AUC of 0.84 (95% CI: 0.77-0.90), a sensitivity of 64.3%, and a specificity of 83%. In the test cohort (Figure 3B), the AUC was 0.70 (95% CI: 0.48-0.88), with a sensitivity of 50%, and a specificity of 76%.

**Figure 3:**
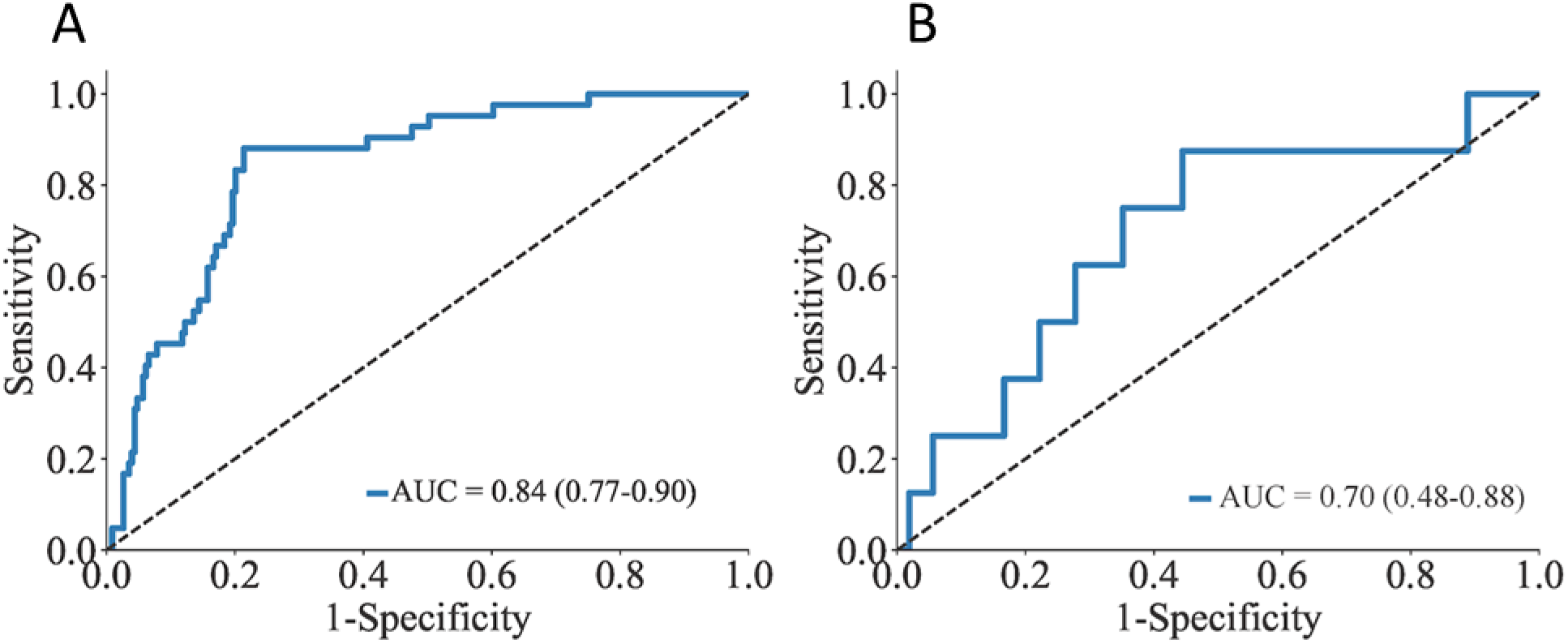
Prediction of High-Risk CAD From ML Model. (A) Training cohort ROC curve for the prediction of high-risk CAD, defined as having either: left main (LM) artery stenosis >50%, 100% stenosis of left anterior descending (LAD), left circumflex (LCx), or right coronary (RCA) artery, or multivessel disease. (B) ROC curve for prediction of high-risk CAD in the test cohort.

### Incremental value of a combined strategy

To investigate the incremental value of ultrasomics in combination with RWMA, a logistic regression model was developed using the ultrasomic prediction probabilities from the primary model, along with the RWMA score. This composite model predicted severe CAD (Figure 4 A-B) with a higher training AUC of 0.82 (95% CI: 0.76-0.88) and test AUC of 0.83 (95% CI: 0.68-0.94) than the corresponding training AUC of 0.67 (95% CI: 0.61-0.72, p<0.0001) and test AUC of 0.70 (95% CI: 0.56-0.81, p=0.037), for RWMA alone. Similarly, for predicting high-risk CAD (Figure 4 C-D), this composite model showed a higher training AUC of 0.86 (95% CI: 0.79-0.92) and a test AUC of 0.76 (95% CI: 0.53-0.96) than the corresponding training AUC of 0.69 (95% CI: 0.63-0.74, p<0.0001) and test AUC of 0.73 (95% CI: 0.60-0.84, p=0.64), for RWMA alone.

**Figure 4:**
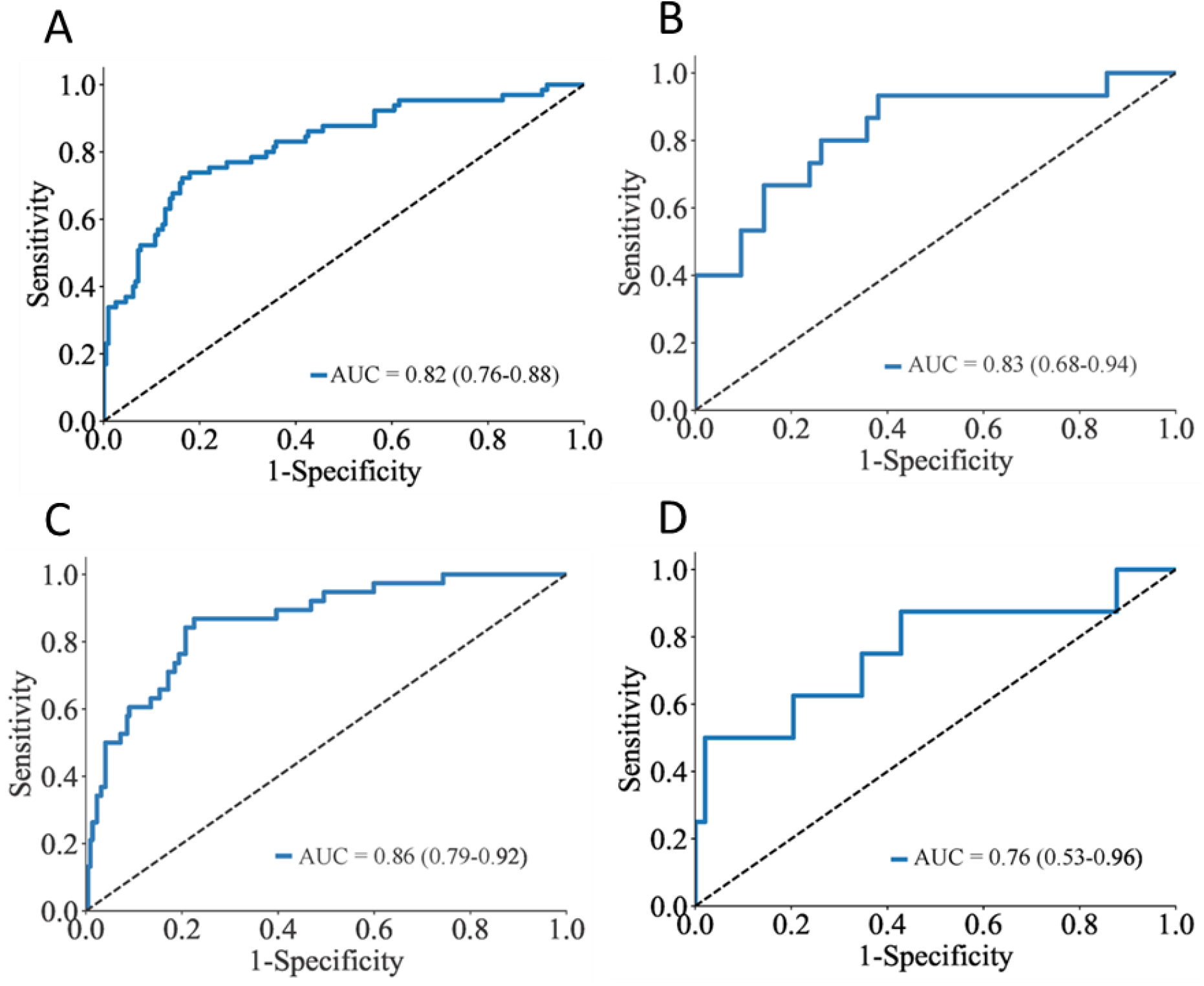
Predicting severe CAD and high-risk CAD using a logistic regression model that combines ultrasomics with regional wall motion abnormalities. ROC curve for predicting severe CAD in the training cohort (A) and the test cohort (B). ROC curve for the prediction of high-risk CAD in the training cohort (C) and the test cohort (D).

## Discussion

To the best of our knowledge, this is the first study that shows the feasibility of an ultrasomics-enabled machine learning model to predict severe obstructive CAD. The ultrasomics model was developed in a heterogeneous cohort of patients from three different institutions using resting echocardiography views (PLAX and A4C views). Despite the lack of any clinical cardiovascular risk factors, the model showed robust performance for predicting severe stenosis in major coronary vessel territories (>70%) and high-risk CAD (left main stenosis >50%, multivessel stenosis >70%, or 100% occlusion). Furthermore, the present study demonstrated that the combination of ultrasomics with RWMA analysis improved the performance of resting echocardiography analysis for predicting both severe and high-risk CAD.

Our previous work in an animal model and in patients undergoing cardiac magnetic resonance imaging had shown the potential of ultrasomics in identifying acoustic reflection patterns related to changes in myocardial architecture that are not identifiable by visual analysis (9,15). The present investigation extends this work by applying ultrasomics to clinical scenarios. Interestingly, obstructive CAD is associated with changes in myocardial architecture characterized by progressive myocyte hypertrophy and interstitial fibrosis, related to underlying risk factors, ischemia, and infarction (16,17). We speculate that ultrasomics may assess obstructive CAD through the quantification of subtle changes in myocardial architecture and textural patterning seen in patients with obstructive CAD.

Given the heterogeneous presentations arising from varying degrees of disease, an individualized approach is needed for risk assessment and interventions. While it is well recognized that even patients with non-obstructive CAD are at an increased risk for future adverse cardiac events, the choice of interventions obviously differs with the presence of severe obstructive CAD; these patients require timely revascularization. Clinical guidelines have therefore recommended the use of CCTA as a first-line diagnostic option in symptomatic persons with a low to intermediate pretest probability of having obstructive CAD (18). However, as seen in our study, a considerable number of people who undergo CCTA have mild or no CAD. Thus, there is a growing interest in developing AI-based approaches to improve patient selection aiming to enhance cost-effectiveness and diagnostic yield of CCTA, as AI-augmented imaging raises physician diagnostic accuracy and can detect more subtle signs indicative of CAD (19–21).

Since cardiac ultrasound is frequently performed in patients with suspected CAD, ML is being increasingly utilized for the quantification of global and regional LV function (22). However, the literature is limited to the prediction of obstructive CAD from echocardiography images beyond gross hemodynamic and structural changes. Upton et al. used ML models built from stress echocardiograms and trained them using an ensemble machine-learning approach to identify patients with severe CAD. Using 31 novel features that were combined into their ML model, the authors were able to achieve an AUC of 0.934 with a high specificity of 85.7% (23). Using a combination of ultrasomics and RWMA from resting studies, our model demonstrated improved predictive value for both severe and high-risk CAD, with a specificity greater than 94% for both endpoints. Furthermore, our model’s applicability to the bedside using limited echocardiography views makes it more feasible for use by clinicians at the point-of-care. Thus, an ultrasomics model implemented at the point-of-care has relevance towards the development of diagnostic tools that may aid appropriate downstream testing. Further evaluation in prospective studies will be of value to understand the impact of new predictive models and their incorporation within clinical decision support systems.

The ultrasomics model may also be beneficial for high-risk patients presenting with unstable coronary syndromes. Recent studies demonstrate that nearly 1/3^rd^ of patients with NSTEMI have high-risk coronary anatomies including an acute occlusion of their culprit artery on invasive coronary angiography (24,25). These patients have double the size of infarction and display significantly higher major adverse cardiac event rates and all-cause mortality than those without total occlusion (26,27). Since the ultrasomics model in the present study identified patients with high-risk CAD, including those with left main disease, multivessel disease, and 100% coronary occlusion, future studies should prospectively assess the incremental value of ultrasomics models in patients with ACS - specifically for identifying patients with occult coronary occlusion who present with NSTEMI and develop large infarcts in the absence of timely interventions.

### Limitations and Future Directions

We analyzed a heterogeneous group of patients from multiple institutions with varying pretest probabilities of CAD. This resulted in class imbalance with the CCTA cohort having a lower prevalence of obstructive CAD than the invasive group. Moreover, the number of patients enrolled at each site was small and we used a strategy of temporal stratification to develop training and test cohorts to provide a realistic evaluation. The developed model would need to be tested in a prospective study where patients are enrolled with uniform inclusion and exclusion criteria. The highest yield would be an assessment performed in patients who present with chest pain in acute care settings and emergency departments where there is an immediate need to identify and efficiently triage patients with high-risk obstructive CAD.

The integration of radiomic-based ML tools for clinical translation, including echocardiography, remains complex but within reach. Most importantly, standardizing and automating the process for ultrasomics extraction and segmentation of myocardium within echocardiography is required for consistently valid model formation (28). Strict standardization of workflow and harmonization of methods are necessary in the ultrasomics pipeline. Integration of models within echocardiography carts to improve diagnostic validity can circumvent scanner-to-scanner variability that can influence results of ultrasomics extraction, image-processing techniques, feature-level optimization, and batch-prediction normalization (29). On the other hand, the use of combined handcrafted ultrasomics with deep ultrasomics may improve model generalizability and its ability to be placed in cloud-based platforms. These implementation strategies would need to be tested in future investigations.

## Conclusion

We demonstrated the ability of ML-enhanced predictive models using ultrasomics to predict severe, obstructive CAD and high-risk CAD in patients more accurately than the traditional assessment of RWMA alone. Our model was developed using limited echocardiography views and requires further external validation specifically when used at the bedside using POCUS as a cost-effective AI tool for screening for obstructive CAD.

## Data Availability

All data produced in the present study are available upon reasonable request to the authors.

